# Antiestrogenic therapy effect on brain structure and mental health in women with breast cancer: a longitudinal MRI study

**DOI:** 10.64898/2026.08.14.26360067

**Authors:** Serenay Yazıcı Sarıkaya, Berfin Gülbahçe, Ann-Christin S. Kimmig, Sara Y. Brucker, Benjamin Bender, Uta Hoopmann, Markus Hahn, Anna Wikman, Birgit Derntl

**Author notes:** Shared first authorship: Serenay Yazıcı Sarıkaya and Berfin Gülbahçe. Corresponding Author: Department of Psychiatry and Psychotherapy, Tübingen Center for Mental Health (TüCMH), University of Tübingen, Tübingen Germany. Email addresses.

## Abstract

Antiestrogenic therapy is widely used in the treatment of hormone receptor–positive breast cancer and alters estrogen signaling through different mechanisms, which may affect brain regions sensitive to estrogenic modulation. However, its early effects on brain architecture remain poorly understood. In this study, we examined whether the initiation of antiestrogenic therapy (tamoxifen or letrozole) is associated with short-term changes in brain structure and psychological symptoms, and whether these changes differ between the two treatment types. For now, data from twenty women with breast cancer and twenty healthy controls undergoing MRI scanning and psychological assessments at baseline (t1) and again approximately 2–3 weeks later (t2) were used. Patients started antiestrogen therapy immediately after the first assessment. Structural analyses included whole-brain cortical thickness and gyrification, as well as region-of-interest measures of hippocampal and amygdala volume. Changes in psychological parameters were also assessed, and hormone levels were measured but are not reported here.

No robust time-by-group effects were observed for total brain volume, cortical thickness, gyrification, or hippocampal and amygdala volume after correction for multiple comparisons. An exploratory within-patient analysis identified a localized increase in cortical thickness in the right anterior insula/inferior frontal operculum; however, the corresponding time-by-group interaction was not significant. Somatic depressive symptom scores showed a significant time-by-group interaction, with scores increasing in the breast cancer group but remaining stable in healthy controls. Across time points, women with breast cancer also reported higher overall depressive symptoms and state anxiety and lower positive affect than healthy controls. Exploratory associations between changes in brain structure and psychological symptoms were observed at uncorrected thresholds but did not survive correction for multiple comparisons.

In this interim sample, no robust group-level macrostructural brain changes were detected over the first 2–3 weeks following initiation of antiestrogen therapy. However, this does not exclude the possibility of early structural effects, which may be subtle or heterogeneous and therefore difficult to detect in the current sample. Somatic depressive symptoms increased in the BC group relative to healthy controls during this early treatment period, while exploratory neural findings suggested potential localized changes and individual-difference associations that warrant cautious interpretation and require confirmation in larger samples. Recruitment is ongoing toward the prospectively defined final sample.

## 1. Introduction

Breast cancer (BC) is the most frequently diagnosed cancer in women worldwide and represents a major public health concern. In 2020, it accounted for nearly one-quarter of all new cancer cases among women (1–4). Improvements in early detection and treatment have substantially increased survival rates, leading to a growing number of long-term survivors. As a result, more recent work has focused on the broader consequences of BC and its treatment, including potential effects on cognitive changes, quality of life (QoL), and mental health (5–7).

Estrogen receptors are widely distributed across the brain, including regions such as the amygdala and hippocampus that are central to emotion regulation, stress reactivity, and memory (8). Via binding to the receptors, estrogens are also known to exert neuroprotective effects through the regulation of neurotransmitter systems, synaptic plasticity, and stress-related pathways, which are important for maintaining cognitive performance and emotional stability (9–11). Withdrawal and decline of estrogen levels, whether occurring naturally during menopausal transition or induced pharmacologically through antiestrogen therapy, may therefore influence emotional and cognitive functioning (12).

Antiestrogen therapy represents a cornerstone in the treatment of hormone receptor–positive BC, which accounts for the majority of BC cases (13,14). Common therapeutic strategies include selective estrogen receptor modulators (SERMs), aromatase inhibitors, and ovarian suppression. Tamoxifen, the most extensively studied SERM, has been widely used for several decades and remains a standard treatment, particularly for premenopausal women (15). Aromatase inhibitors such as letrozole reduce estrogen production by inhibiting aromatase, the enzyme responsible for the peripheral conversion of androgens into estrogens in postmenopausal women (16,17).

Although antiestrogen therapy substantially improves survival outcomes in BC, several studies showed that treatment-related side effects and the broader physical and psychological burden of the disease negatively affect QoL, body image, and mental health in affected women (18–31). Since these treatments interfere with estrogen production or synthesis, their effects may be particularly relevant in brain regions with high estrogen receptor density. However, the impact of antiestrogen therapy on brain structure remains unclear. A recent meta-analysis shows that most neuroimaging studies in BC have focused on chemotherapy-related changes (“chemobrain”) (Godaetr & Drame, 2025), while the effects of antiestrogen therapy have received comparatively little attention.

The few studies directly investigating the effect of antiestrogen therapy on brain structure reported reductions in gray matter volume (GMV) and cortical thickness in frontal and temporal regions, including the hippocampus: A cross-sectional study examined the effects of tamoxifen (TAM) on brain structure and cognitive function in postmenopausal women diagnosed with BC (32). The study compared women receiving TAM (n=10) for two years with two healthy groups: women currently using menopausal hormone therapy and taking estrogens (ERT+, n=15) and women not receiving estrogens (ERT−, n=15). The TAM group showed significantly smaller right hippocampal volumes compared with the ERT+ group. This structural difference was further associated with impaired semantic memory performance in the TAM group, suggesting a potential negative effect of TAM on hippocampal-dependent cognitive processes relative to estrogen exposure. In addition, a longitudinal study of postmenopausal BC patients (n=29) treated with letrozole (LET) for one year reported GMV reductions in the hippocampus and amygdala, as well as decreased cortical thickness in the prefrontal, parietal, and insular cortices, after one year of therapy (33). However, evidence on the early effects of antiestrogen therapy on brain structure remains limited.

Thus far, the two previously published structural imaging studies have primarily relied on volumetric and cortical thickness measures. To our knowledge, no study has investigated gyrification measures. Therefore, this ongoing study examined whether the initiation of antiestrogen therapy is associated with early changes in brain structure, focusing on whole-brain cortical thickness and gyrification as well as region-specific volume and cortical thickness. We selected regions that have previously been shown to show differences between groups with and without antiestrogenic therapy (32), are densely packed with estrogen receptors (34,35), and are essential for stress reactivity, mood regulation, and mental health. Using a longitudinal MRI design, structural brain measures were assessed before treatment initiation (t1) and again two to three weeks later (t2), before breast surgery. In addition, psychological measures were collected at both time points to examine potential associations between structural brain changes and affective symptoms during the early phase of treatment.

Based on previous findings, we expected that antiestrogenic therapy would be associated with reductions in brain volume in areas involved in emotional and cognitive processing, particularly in the hippocampus and amygdala (32,33). Furthermore, we expected that patients after starting antiestrogen therapy will report increased affective symptoms and reduced well-being during this early treatment period (36–38). Additionally, we predicted that treatment-related structural brain changes would be associated with changes in psychological distress, such that higher levels of distress would correspond to lower cortical and subcortical indices. Finally, exploratory analyses repeated these analyses separately within antiestrogenic subgroups (e.g., tamoxifen vs. letrozole) to examine whether the observed psychological and structural changes differed by treatment type.

## 2. Methods

### 2.1 Participants

For now, 49 female participants (29 patients, 20 healthy controls) were initially recruited. Recruitment is continuing toward a prospectively defined target sample of N = 80. The target sample size was determined independently of the interim results. All findings should therefore be considered preliminary and will be updated in a subsequent version. All participants were aged between 43 and 70 years. Eligibility criteria included having a body mass index (BMI) between 18 and 30 kg/m², being a non-smoker, being of European descent, having at least a high school diploma and being fluent in German. The study was approved by the Ethics Committee of the Medical Faculty Tübingen (508/2023BO2, 11.10.2023). All participants provided written informed consent prior to participation.

Patients were recruited at the Women’s Clinic of the University Hospital Tübingen when seeking diagnosis for estrogen-receptor positive BC. The treating gynecologist informed them about the study and patients were enrolled after being diagnosed before the first intake of antiestrogen treatment. All patients were enrolled in the study only if they did not have any major exclusion criteria, including giving birth or breastfeeding within the last year, male breast cancer, ongoing chemotherapy treatment, cancer originating from any organ other than the breast, or severe medical conditions such as diabetes or stroke. Exclusion criteria for all participants included the presence of neurological or mental disorders, as assessed using the Structured Clinical Interview for DSM-5, Clinical Version (SCID-5-CV) (39). Participants were also excluded if they were not MRI-compatible (e.g., active implants), if they had medical conditions other than BC (including endocrine, metabolic or chronic diseases), if they used medications known to affect brain function (including hormonal contraception), if they reported substance abuse, if they had a history of sexual trauma or abuse, or if they declined to provide informed consent.

### 2.2 Study Design and Materials

Participants completed two MRI sessions (t1 and t2). The assessment protocol was the same for both sessions and included an anatomical scan, a resting-state scan, diffusion tensor imaging (DTI), a functional paradigm assessing reward processing, i.e. the effort allocation task (40), a blood sample, and a set of questionnaires. During t1, which occurred before the initiation of antiestrogen therapy, participants completed a set of questionnaires covering topics such as body image, emotion regulation, relationship satisfaction, and quality of life. This was followed by the MRI scan. t2 took place two to three weeks after t1. Patients with BC started antiestrogen therapy (tamoxifen or letrozole) immediately after t1. This allowed us to examine changes early on following treatment initiation, before surgical intervention (see Figure 1).

**Figure 1.**
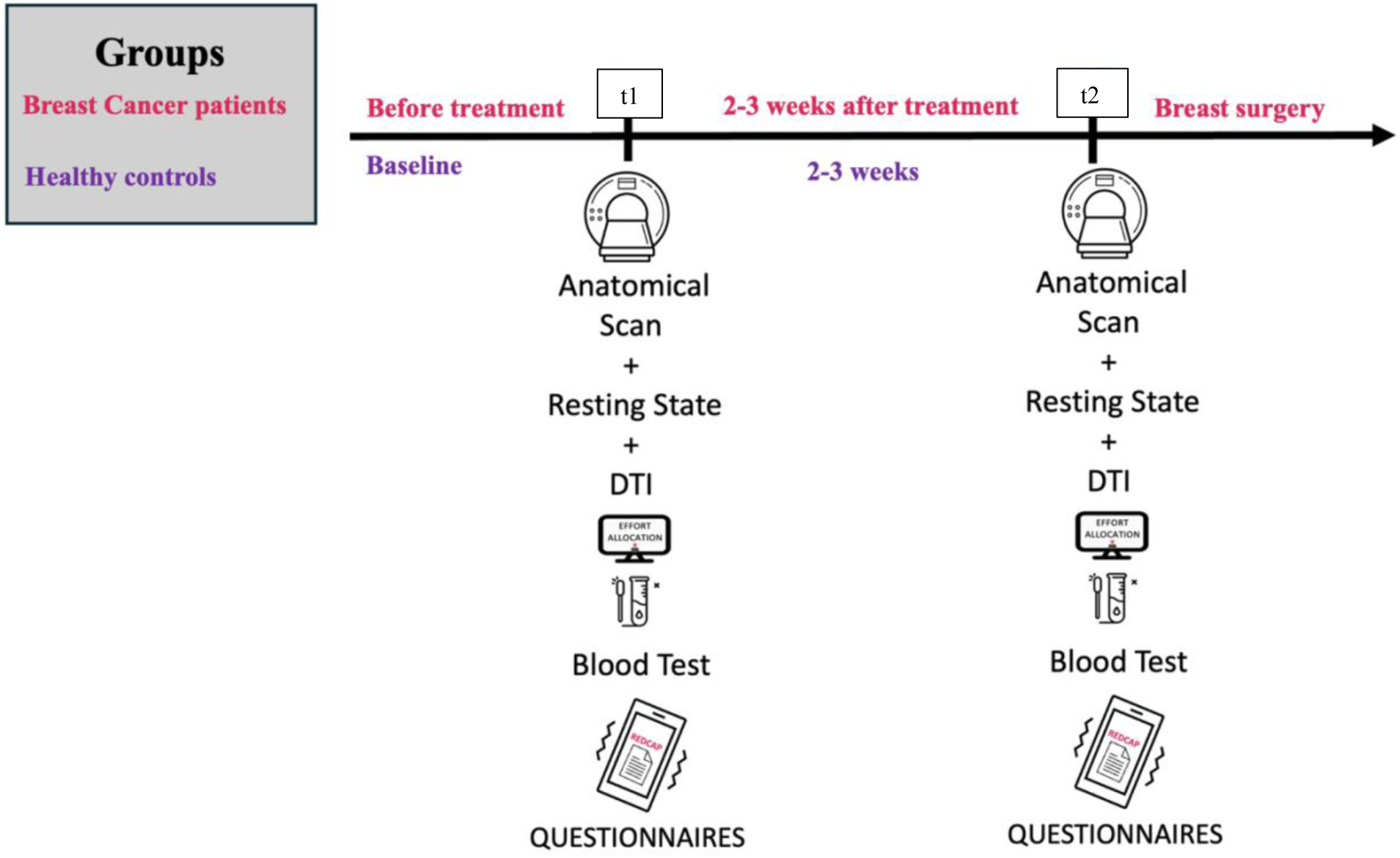
Study design and timeline of the longitudinal MRI protocol. Two MRI sessions were conducted: t1 and t2. After the t1 session, breast cancer patients-initiated antiestrogen therapy (tamoxifen or letrozole), and the t2 session was conducted two to three weeks later. The MRI sessions included an anatomical scan, a resting state scan, and an effort allocation scan.

#### Psychological Outcomes

Depressive symptoms were assessed using the Beck Depression Inventory-II (BDI-II, 37), a 21-item self-report questionnaire measuring the severity of depressive symptoms over the past two weeks. To assess more nuanced group and time differences, the somatic and cognitive-affective subscales of BDI-II were also used separately (42). Affective states were measured with the Positive and Negative Affect Schedule (PANAS, 39), which consists of two 10-item subscales assessing positive and negative affect. State anxiety was assessed using the State-Trait Anxiety Inventory – State version (STAI-S) (Laux et al., 1981), a 20-item self-report scale measuring current levels of anxiety. Higher scores on each scale indicate greater levels of the respective psychological construct.

### 2.3 MRI Data Acquisition and Processing

#### Data acquisition

MRI data were collected on a 3T Siemens Prisma scanner (Siemens, Erlangen, Germany) located at the Department of Psychiatry and Psychotherapy, University Hospital Tübingen. Using a 64-channel head coil, high-resolution anatomical images were acquired using a T1-weighted magnetization-prepared rapid gradient echo sequence (MPRAGE; TR = 2400 ms, TE = 2.22 ms, voxel size = 0.8 mm³, flip angle = 8°, field of view = 256 × 256 mm, 208 sagittal slices, acceleration factor PE = 2). The acquisition time was approximately 10 minutes.

#### Structural MRI Data Processing

Structural MRI data were processed using the CAT12 toolbox (version 12.9; https://neuro-jena.github.io/cat/) implemented in SPM12 (v7771; https://www.fil.ion.ucl.ac.uk/spm) within MATLAB (version 2023b; https://uk.mathworks.com/), following the procedures described previously (44). Briefly, a longitudinal processing pipeline was applied to estimate surface-based morphometry measures, including cortical thickness, Toro’s gyrification index, sulcal depth, and total brain volume (TBV). Preprocessing included correction for magnetic field inhomogeneities, skull stripping, and removal of the cerebellum and brainstem. Images were then segmented into grey matter (GM), white matter, and cerebrospinal fluid (CSF). Cortical surfaces were reconstructed using the projection-based thickness method implemented in CAT12 (45). This approach accounts for partial volume effects, sulcal blurring, and sulcal asymmetries without requiring explicit sulcus reconstruction. Surface processing further included topology correction and spherical mapping (43). To enable interindividual comparability, individual cortical surfaces were spatially normalized to the fsaverage template using spherical mapping with minimal distortions.

In addition to cortical thickness, the local gyrification index was calculated based on the ratio of cortical surface area reflecting local cortical folding (47). Sulcal depth was estimated as the Euclidean distance between the central cortical surface and its convex hull and subsequently transformed using a square-root function to improve normality. To enable interindividual comparability, individual cortical surfaces were spatially normalized to the fsaverage template using spherical mapping with minimal distortions. Prior to statistical analysis, normalized surfaces were smoothed using a 20-mm full-width at half-maximum (FWHM) kernel for gyrification and sulcal depth measures and a 12-mm FWHM kernel for cortical thickness (48–50).

In addition to surface-based measures, region-of-interest (ROI) analyses were conducted focusing on the hippocampus and amygdala. T1 images were subsequently normalized to Montreal Neurological Institute (MNI) space with a voxel size of 0.8 mm. ROI masks were created based on the Neuromorphometrics atlas using the Image Calculator tool in SPM12. GM was extracted for the left and right hippocampus and amygdala using the get_totals script (owned by G. Ridgeway) as previously described by Pletzer and colleagues (51). Since all baseline (t1) volumes were strongly correlated with t2 volumes (p < .001), total intracranial volume (TIV) correction was based on baseline estimates to account for interindividual differences in head size following De Paepe et al. (52).

### 2.4 Statistical Analysis

Sociodemographic characteristics of the BC vs HC were compared using independent samples t-test for continuous variables (age) and chi-square test for categorical variables (education level, family situation, marital status, economic status).

The primary analyses examined longitudinal changes in structural brain measures. Repeated-measures analyses of variance (ANOVAs) were conducted for volumetric and surface-based measures, with session (t1 vs. t2) as the within-subject factor and group (BC vs. HC) as the between-subject factor. Age was included as a covariate in all models. Whole-brain voxel-wise GMV analyses were conducted using CAT12. Statistical maps were initially thresholded at *p* < .001, uncorrected, and correction for multiple comparisons was subsequently applied using false discovery rate (FDR) correction. Within the BC group, longitudinal whole-brain voxel-based morphometry changes between t1 and t2 were additionally examined using paired-samples *t* tests. Region-of-interest analyses of left and right hippocampal and amygdala GMVs were examined using repeated-measures ANOVAs with the same within- and between-subject factors. Cortical thickness and gyrification were also assessed longitudinally using repeated-measures models.

Secondary analyses examined longitudinal changes in psychological outcomes. Separate repeated-measures ANOVAs were conducted for total depressive symptoms, somatic and cognitive-affective depressive symptoms, state anxiety, and positive and negative affect. Session was entered as the within-subject factor, group as the between-subject factor, and age as a covariate. Where a significant time x group interaction was observed, follow-up comparisons were conducted to characterize the interaction.

Brain–behavior associations were examined exploratorily within the BC group after the primary structural and secondary psychological analyses. Change scores (Δ = t2 − t1) were calculated for left and right hippocampal and amygdala GMVs, total depressive symptoms, somatic and cognitive-affective depressive symptoms, state anxiety, and positive and negative affect. Only the volumetric data were log-transformed to reduce skewness. Associations between changes in regional GMV and changes in psychological outcomes were assessed using age-adjusted Spearman correlations. Each variable was first rank transformed, after which the ranked brain and psychological variables were separately residualized with respect to ranked age using ordinary least squares regression. Pearson correlations between the resulting residuals provided age-adjusted rank-based correlation coefficients. Confidence intervals were estimated using nonparametric bootstrap resampling with 5,000 iterations. FDR correction was applied to account for multiple testing across the brain–behavior correlations.

Exploratory analyses were subsequently conducted within the patient sample to examine potential differences between participants receiving tamoxifen and those receiving letrozole. Repeated-measures ANOVAs included session as the within-subject factor and treatment group as the between-subject factor, with age included as a covariate. Subgroup-specific longitudinal changes in structural and psychological outcomes were also examined separately within the tamoxifen and letrozole groups. Brain–behavior associations identified in the full BC group were additionally examined within each treatment subgroup. Given the small subgroup sample sizes, these analyses were considered exploratory.

Further analyses of the CAT12 output, including ANOVAs, *t* tests, and correlations, were conducted using JASP version 0.19 (50). Results were visualized using Python version 3.12.4. Statistical significance was set at *p* < .05. Effect sizes for significant ANOVA results are reported as partial eta squared (η²ₚ).

## 3. Results

### Sample Description

A total of 49 participants were initially recruited. Following exclusions due to MRI incompatibility (*n* = 6) and incomplete data (*n* = 3), the preliminary sample consists of 20 patients with BC and 20 HC. Groups did not differ significantly in terms of age (56.6 ± 7.2 vs. 53.8 ± 7.6 years, p = .180), education level (p = .132), or family situation (p = .144), indicating they were well-matched. Among patients, eight received tamoxifen and were pre- (n=7) or perimenopausal (n=1), while 12 received letrozole and were mostly postmenopausal (n=11, one patient was perimenopausal). Demographic and clinical characteristics are presented in Table 1. Patients receiving tamoxifen or letrozole did not differ significantly in demographic characteristics, except for age and menopausal stage, with letrozole receiving patients being older and postmenopausal.

**Table 1.** Participant Characteristics.

|  | Breast cancer patients<br>(n=20) | Healthy controls<br>(n=20) |  |
| --- | --- | --- | --- |
| Characteristic | Mean ± SD or n (%) | | p-value |
| **Age** | 56.6± 7.2 | 53.8± 7.6 | .180 |
| **Menopausal Status** |  |  |  |
| Pre-menopausal | 7 (35%) | 8 (40%) |  |
| Peri-menopausal | 2 (10%) |  |  |
| Post-menopausal | 11 (55 %) | 12 (60%) |  |
| **Medication** |  |  |  |
| Tamoxifen | 8 (40%) | n/a |  |
| Letrozole | 12 (60%) | n/a |  |
| **Marital status** |  |  |  |
| Partner | 19 (95%) | 18 (90%) | .144 |
| No partner | 1 (5%) | 2 (10%) |  |
| **Children** |  |  |  |
| Yes | 17 (85%) | 17 (85%) |  |
| No | 3 (15%) | 3 (15%) |  |
| **Education level** |  |  | .132 |
| Less than university degree | 7 (35%) | 9 (45%) |  |
| University degree | 13 (65%) | 11 (55%) |  |

*Note.* Values are presented as mean ± standard deviation or number (percentage). M: mean; SD: standard deviation

### Structural MRI

#### Whole-brain voxel-wise analyses

A mixed-design ANOVA showed no significant main effect of time on whole-brain volume (p = .412), no main effect of group p = .638, and no time x group interaction (p = .731), indicating that the magnitude of pre-to-post change did not differ between the two groups (BC vs HC) (Table 2).

**Table 2.**
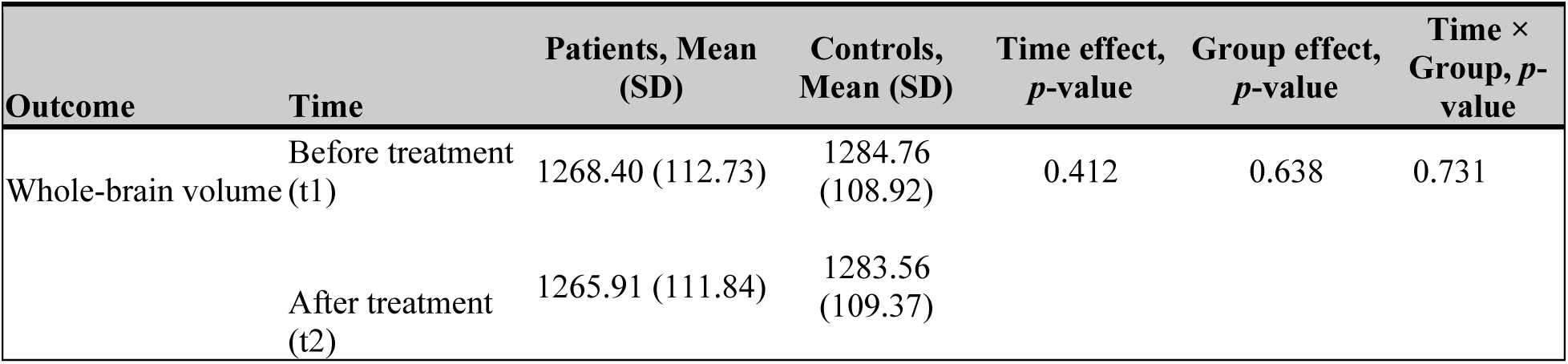
Mixed ANOVA results for whole-brain volume by time (pre-vs. post-assessment), group (patients vs. controls), and the time × group interaction.

| Outcome | Time | Patients, Mean (SD) | Controls, Mean (SD) | Time effect, *p*-value | Group effect, *p*-value | Time × Group, *p*-value |
| --- | --- | --- | --- | --- | --- | --- |
| Whole-brain volume | Before treatment (t1) | 1268.40 (112.73) | 1284.76 (108.92) | 0.412 | 0.638 | 0.731 |
|  | After treatment (t2) | 1265.91 (111.84) | 1283.56 (109.37) |  |  |  |

#### Cortical thickness

No clusters survived cluster-level FWE correction for the main effect of time, the main effect of group, or the time x group interaction (all *p*FWE, cluster > .10). In an exploratory analysis restricted to the BC group, the t2 > t1 contrast identified a cluster encompassing the right insular and inferior frontal opercular regions (*k* = 62, *p* < .001, uncorrected; Figure 3 and Table 3). This cluster did not survive cluster-level FWE correction (*p*FWE, cluster = .191), although the peak vertex survived peak-level FWE correction (*T* = 4.65, *Z* = 4.33, *p*FWE, peak = .035) and FDR correction (*q*FDR, peak = .030; MNI coordinates [31, 5, −2]). No significant clusters were observed for the reverse within-group contrast, t1 > t2 (see Figure 2).

**Figure 2.**
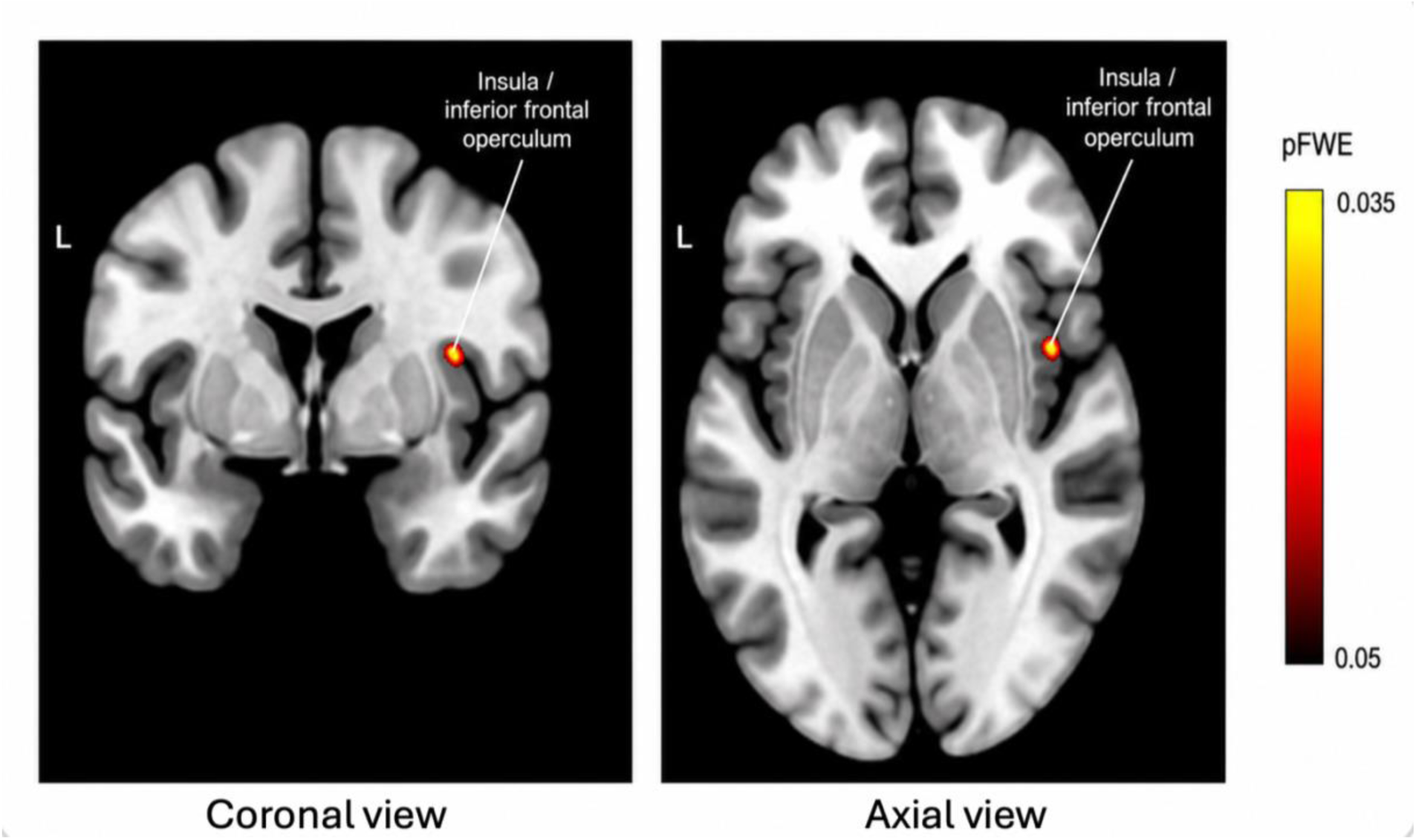
Anatomical localization of the exploratory cortical thickness finding in the breast cancer group. Orthogonal views are centered on the peak MNI coordinate [31, 5, −2] in the right anterior insula/inferior frontal operculum. The marker indicates the location of the peak-level increase in cortical thickness from t1 to t2 within the breast cancer group. The cluster was identified at an exploratory threshold of p < .001, uncorrected, and did not survive correction for multiple comparisons at the cluster level. The figure shows the anatomical location of the peak and not the full spatial extent of the cluster.

**Figure 3.**
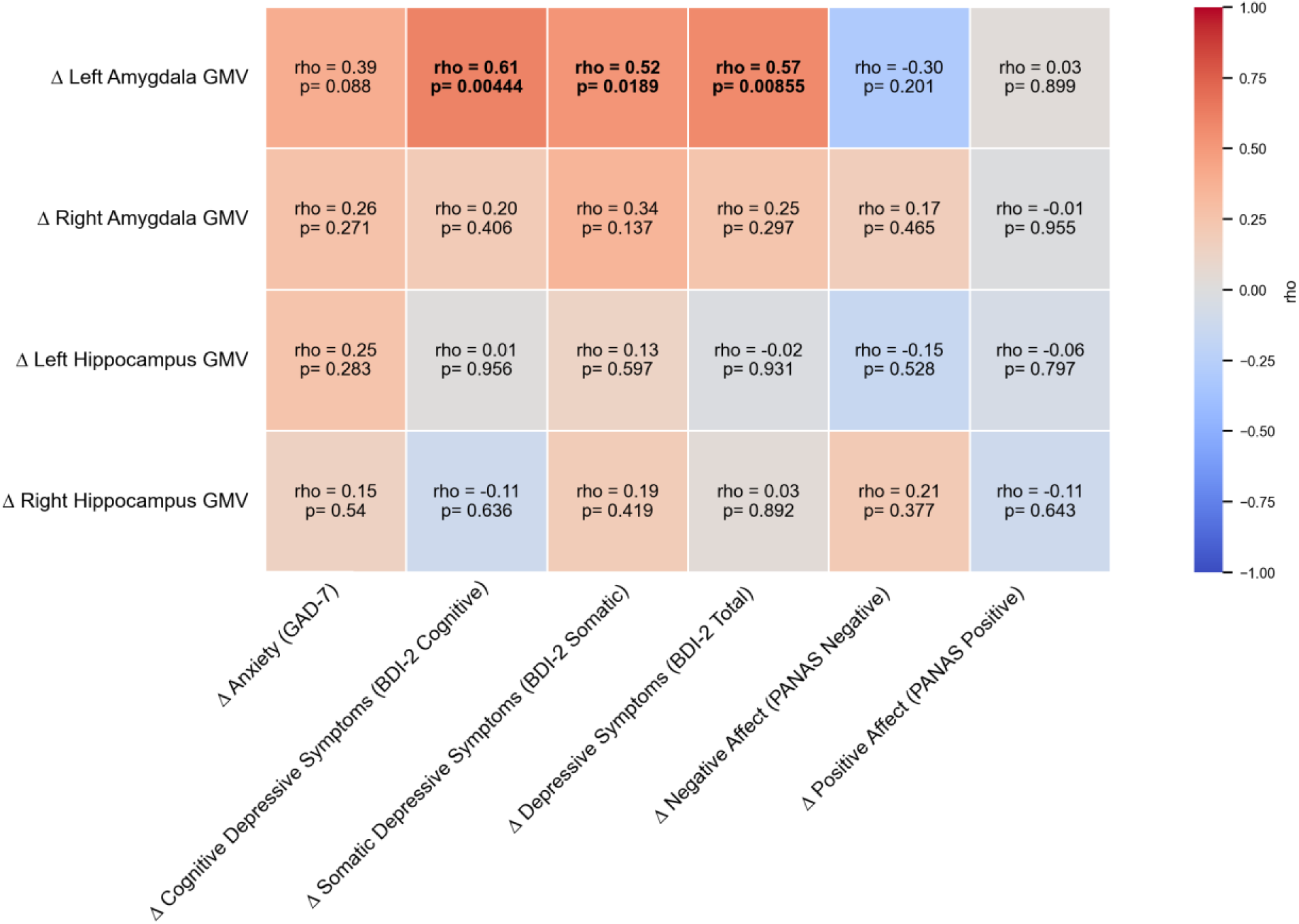
Associations between changes in (GMV) and mental health measures. The heatmap illustrates age-adjusted Spearman correlations between changes in GMVs of hippocampus and amygdala and changes in positive and negative affect, anxiety, cognitive, somatic and total depressive symptom scores in BC group. Positive associations were observed for left amygdala GMV and measures of depressive symptoms. Note. Δ = change score (t2 − t1), GMV: Gray matter volume, BDI-2: Beck Depression Inventory-2.

**Figure 4.**
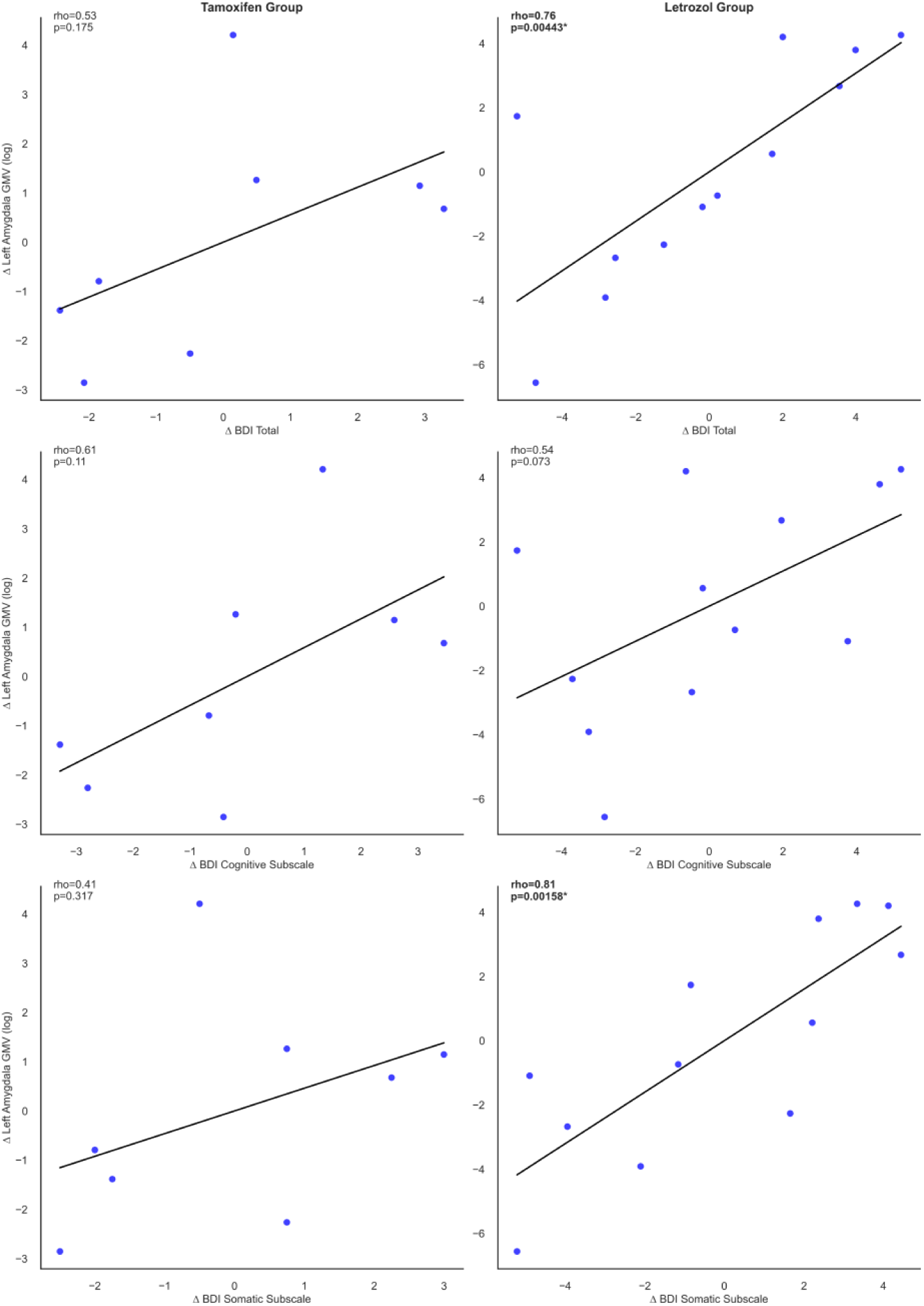
Associations between changes in amygdala GMV and depressive symptoms. Scatterplots illustrating age-adjusted Spearman correlations between changes in left amygdala **GMV** and changes in depressive symptoms (BDI) in the breast cancer group. Separate panels display results for the tamoxifen and letrozole subgroups. Positive associations were observed for **GMV**, particularly in the letrozole group. Note. Δ = change score (t2 − t1). Lines represent fitted regression slopes. GMV: Gray matter volume, BDI-2: Beck Depression Inventory-2.

**Table 3.** Cortical Thickness - Exploratory vertex-level finding.

| Comparison | Region | k | MNI (x, y, z) | pFDR | pFWE |
| --- | --- | --- | --- | --- | --- |
| BC vs. HC | Insula / Inferior Frontal Operculum | 18 | (30, 5, -4) | .907 | .634 |
| BC (t2 > t1) | Right Insula / Inferior Frontal Operculum | 62 | (31, 5, -2) | **.030** | **.035** |
| Letrozole (t2 > t1) | Right Insula / Inferior Frontal Operculum | 62 | (31, 5, -2) | .058 | .142 |

*Note.* Peak-level statistics are reported, as cluster-level correction was not met for any comparison. k = cluster extent (number of vertices); MNI = Montreal Neurological Institute coordinates (x, y, z, in mm). BC = breast cancer; HC = healthy control. Only the BC (t2 > t1) contrast reached peak-level significance after FWE correction ( $p = .035$ ); all other comparisons did not survive correction for multiple comparisons

#### ROI-based volumetric analyses

For amygdala and hippocampus, repeated measures ANOVA revealed no significant main effect of time (all ps > .377, all η²s ≤ 1.905 x 10^-4^) nor group (all ps > .123, all η²s ≤ 0.046), and no significant time x group interactions (all ps > .393, all η²s ≤ 1.784 x 10^-4^) (Table 4). Age was a significant covariate in the group x time analysis of the amygdala and hippocampus GMVs (all ps < .045),(η² = .11–.25), with younger participants exhibiting larger GMV values (Table 4).

**Table 4.** Repeated-Measures ANOVA Results for ROI Volumes (BC vs HC)

|  |  |  |  | Within Subjects Effects |  |  |  |  |  | Between Subjects Effects |  |  |  |
| --- | --- | --- | --- | --- | --- | --- | --- | --- | --- | --- | --- | --- | --- |
| Brain Region and Tissue Type | Time | BC Mean (SD) | HC Mean (SD) | Time p-value | Time $\eta^2$ | Time x Group p-value | Time x Group $\eta^2$ | Time x Age p-value | Time x Age $\eta^2$ | Age p-value | Age $\eta^2$ | Group p-value | Group $\eta^2$ |
| AL GM | t1 | -7.43<br>(0.05) | -7.40<br>(0.09) | .607 | 1.178x<br>10<sup>-4</sup> | .688 | 7.178 x<br>10<sup>-5</sup> | .632 | 1.023x10<sup>-4</sup> | **.045** | 0.101 | .462 | 0.013 |
|  | t2 | -7.43<br>(0.06) | -7.40<br>(0.09) |  |  |  |  |  |  |  |  |  |  |
| AR GM | t1 | -7.49<br>(0.05) | -7.46<br>(0.09) | .629 | 5.583x<br>10<sup>-5</sup> | .732 | 2.791 x<br>10<sup>-5</sup> | .619 | 5.932x10<sup>-5</sup> | **.008** | 0.169 | .434 | 0.014 |
|  | t2 | -7.49<br>(0.05) | -7.46<br>(0.09) |  |  |  |  |  |  |  |  |  |  |
| HL GM | t1 | -6.39<br>(0.06) | -6.34<br>(0.08) | .377 | 1.905x<br>10<sup>-4</sup> | .393 | 1.784 x<br>10<sup>-4</sup> | .435 | 1.485x10<sup>-4</sup> | **.003** | 0.201 | .135 | 0.047 |
|  | t2 | -6.38<br>(0.06) | -6.34<br>(0.08) |  |  |  |  |  |  |  |  |  |  |
| HR GM | t1 | -6.32<br>(0.05) | -6.27<br>(0.09) | .602 | 4.113x<br>10<sup>-5</sup> | .453 | 2.522 x<br>10<sup>-5</sup> | .629 | 3.530x10<sup>-5</sup> | **< .001** | 0.258 | .123 | 0.046 |
|  | t2 | -6.32<br>(0.05) | -6.27<br>(0.08) |  |  |  |  |  |  |  |  |  |  |

*Note.* df= (1, 37) AL: Left Amygdala, AR: Right Amygdala, HL: Left Hippocampus, HR: Right Hippocampus, GM: Gray Matter.

### Psychological outcomes

#### Depressive symptoms

For the total BDI-II score, a main effect of group (F(1,37) = 13.36, p = .016, η² = .134) emerged, with BC reporting higher levels than HC. No time effect (F(1,37) = .015, p = .902) or time x group interaction (F(1,37) = 1.609, p = .213) was observed (Table 5).

**Table 5.**
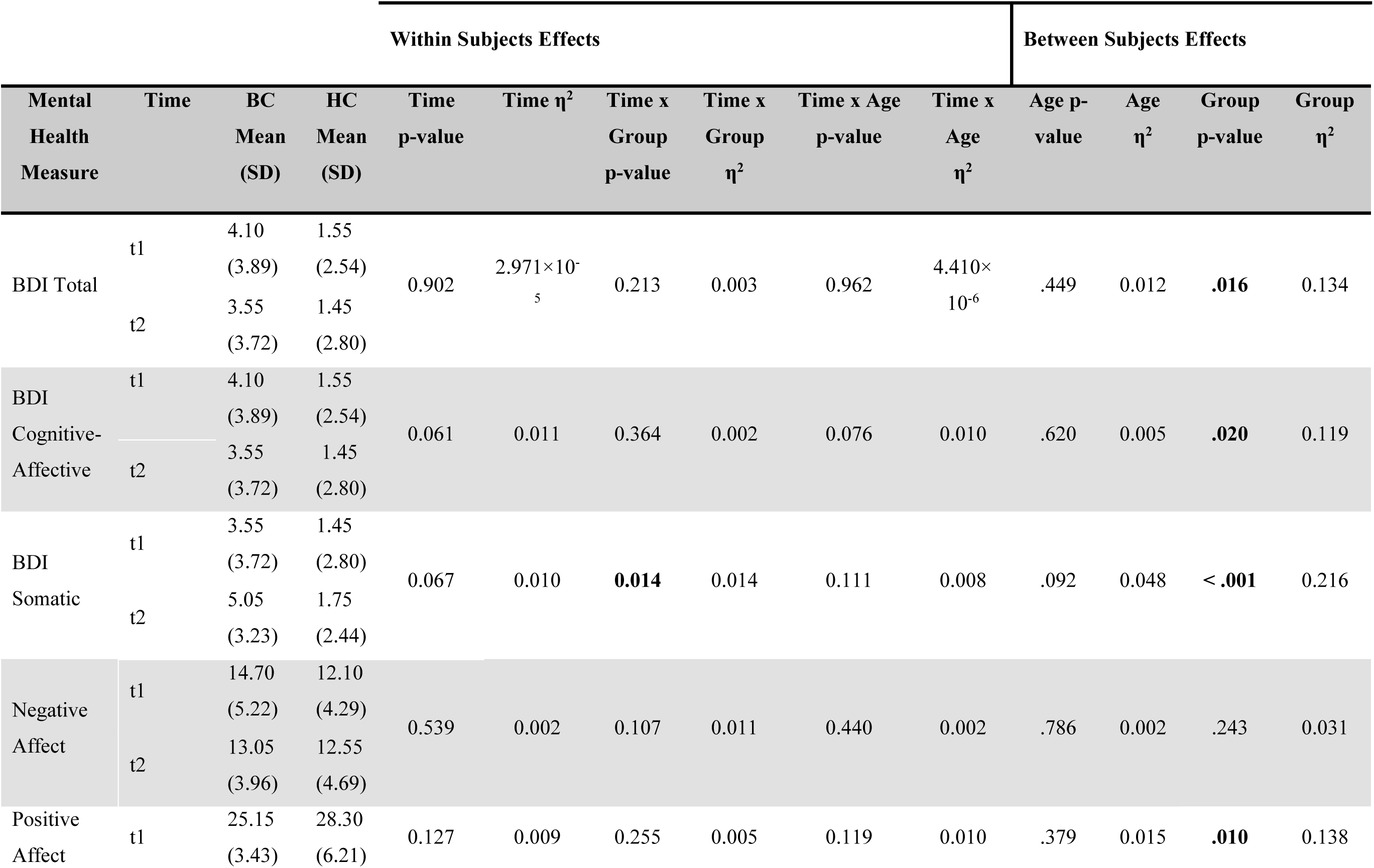
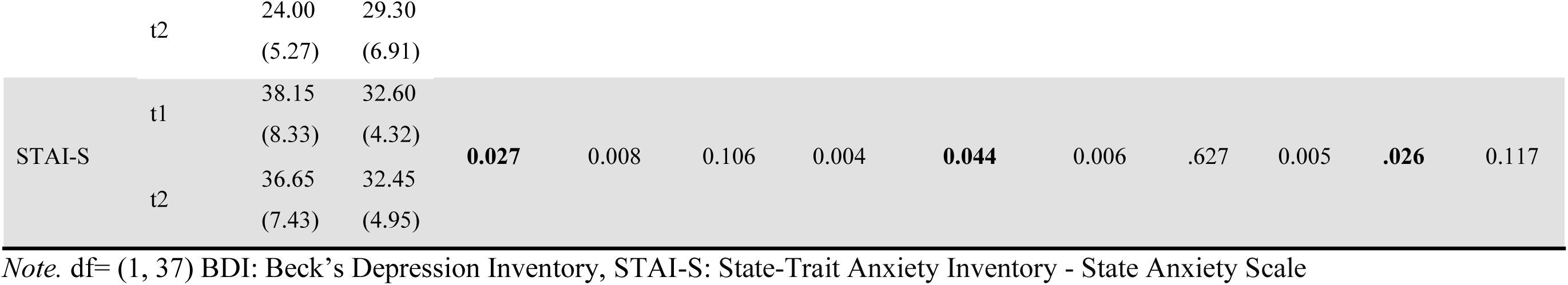
Repeated-Measures ANOVA Results for Psychological Outcomes.

For somatic depressive symptoms, a main effect of group (F(1,37) = 13.36, p < .001, η² = .216), with BC reporting higher levels, and a significant time x group interaction (F(1,37) = 5.05, p = .031, η² = .014) emerged. Time showed a trend, but no significant main effect (F(1,37) = 3.55, p = .067). Disentangling the significant interaction, post-hoc analyses indicated that BC group showed overall higher levels of somatic depression (p < .001). Within group comparison revealed a significant increase only in the BC group (t(19) = -2.837, p = .011, Cohen’s d = -.634) (for mean and SD values see Table 5).

For cognitive depressive symptoms, a main effect of group (F(1,37) = 5.89, p = .020, η² = .119) emerged, with BC reporting higher levels than HC. No time (F(1,37) = 3.73, p = .061) or time x group interaction (F(1,37) = .844, p = .364) was observed (Table 5).

#### State anxiety

A significant main effect of time (F(1,37) = 5.27, p = .027, η² = .008), with overall higher scores at t1, and a main effect of group (F(1,37) = 5.36, p = .026, η² = .117), with higher scores in BC than HC, emerged. The time x group interaction did not reach significance (F(1,37) = .106). Also, a significant time x age interaction emerged in the covariate analysis (F(1, 37) = 4.33, p = .044, η² = .006) (Table 5).

#### Positive affect

A significant main effect of group (F(1, 37) = 7.84, p = .010, η²ₚ = .138) was observed, reflecting lower positive affect in the BC group relative to the HC group. No time (F(1,37) = 2.44, p= .127) or time x group interaction (F(1,37) = 1.33, p = .255) was observed (Table 5).

#### Negative affect

No significant main effect of time (F(1, 37) = .384, p = .539) or group (F(1, 37) = 1.40, p = .243) nor time x group interaction (F(1, 37) = 2.72, p =.107) was observed (Table 5).

### Brain–behavior associations

Exploratory Spearman’s correlation analyses examined associations between changes in brain structure and psychological measures within the BC group (see Figure 3). No results remained significant when corrected for multiple comparisons (all *p-FDR*s > .20). Uncorrected results showed that changes in left amygdala GMV were positively associated with changes in depressive symptoms (ρ = .57, *p* = .009), somatic depressive symptoms (ρ = .52, *p* = .019) and cognitive depressive symptoms (ρ = .61, *p* = .004) (see Table 6a).

**Table 6a.**
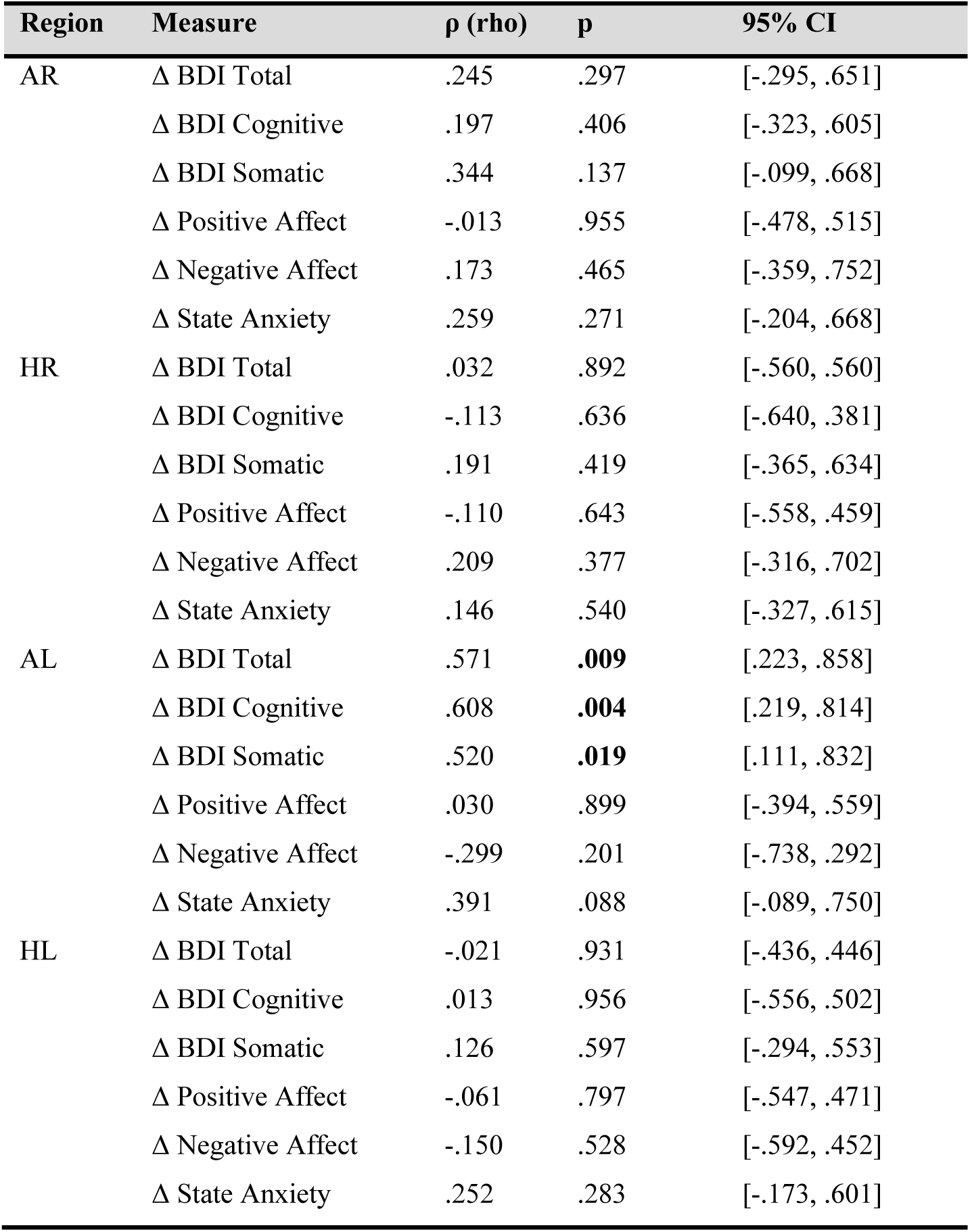
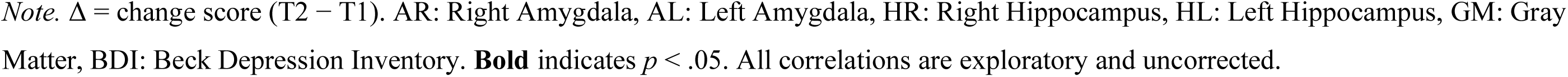
Gray Matter Volume-Behavior Associations in Change Scores (BC Group)

| Region | Measure | $\rho$ (rho) | p | 95% CI |
| --- | --- | --- | --- | --- |
| AR | $\Delta$ BDI Total | .245 | .297 | [-.295, .651] |
| | $\Delta$ BDI Cognitive | .197 | .406 | [-.323, .605] |
| | $\Delta$ BDI Somatic | .344 | .137 | [-.099, .668] |
| | $\Delta$ Positive Affect | -.013 | .955 | [-.478, .515] |
| | $\Delta$ Negative Affect | .173 | .465 | [-.359, .752] |
| | $\Delta$ State Anxiety | .259 | .271 | [-.204, .668] |
| HR | $\Delta$ BDI Total | .032 | .892 | [-.560, .560] |
| | $\Delta$ BDI Cognitive | -.113 | .636 | [-.640, .381] |
| | $\Delta$ BDI Somatic | .191 | .419 | [-.365, .634] |
| | $\Delta$ Positive Affect | -.110 | .643 | [-.558, .459] |
| | $\Delta$ Negative Affect | .209 | .377 | [-.316, .702] |
| | $\Delta$ State Anxiety | .146 | .540 | [-.327, .615] |
| AL | $\Delta$ BDI Total | .571 | **.009** | [.223, .858] |
| | $\Delta$ BDI Cognitive | .608 | **.004** | [.219, .814] |
| | $\Delta$ BDI Somatic | .520 | **.019** | [.111, .832] |
| | $\Delta$ Positive Affect | .030 | .899 | [-.394, .559] |
| | $\Delta$ Negative Affect | -.299 | .201 | [-.738, .292] |
| | $\Delta$ State Anxiety | .391 | .088 | [-.089, .750] |
| HL | $\Delta$ BDI Total | -.021 | .931 | [-.436, .446] |
| | $\Delta$ BDI Cognitive | .013 | .956 | [-.556, .502] |
| | $\Delta$ BDI Somatic | .126 | .597 | [-.294, .553] |
| | $\Delta$ Positive Affect | -.061 | .797 | [-.547, .471] |
| | $\Delta$ Negative Affect | -.150 | .528 | [-.592, .452] |
| | $\Delta$ State Anxiety | .252 | .283 | [-.173, .601] |

*Note.* $\Delta$ = change score (T2 – T1). AR: Right Amygdala, AL: Left Amygdala, HR: Right Hippocampus, HL: Left Hippocampus, GM: Gray Matter, BDI: Beck Depression Inventory. **Bold** indicates $p < .05$ . All correlations are exploratory and uncorrected.

**Table 6b.** Spearman correlations between changes in left amygdala volume and depressive symptoms by treatment group (age-corrected)

| Group | Tissue | Measure | $\rho$ (rho) | p | 95% CI |
| --- | --- | --- | --- | --- | --- |
| Letrozole | GM | $\Delta$ BDI Somatic | **0.80** | **.001** | [.289, .953] |
| Letrozole | GM | $\Delta$ BDI Total | **0.76** | **.004** | [0.03, 1] |
| Letrozole | GM | $\Delta$ BDI Cognitive | 0.54 | .073 | [-.195, .911] |
| Tamoxifen | GM | $\Delta$ BDI Cognitive | 0.60 | .110 | [-.238, .979] |
| Tamoxifen | GM | $\Delta$ BDI Total | 0.53 | .175 | [-.301, .992] |
| Tamoxifen | GM | $\Delta$ BDI Somatic | 0.40 | .317 | [-.453, .994] |

*Note.* $\Delta$ = change score (T2 – T1). GM: Gray Matter. Results are presented separately for the letrozole and tamoxifen groups. $p < .05$ . All correlations are exploratory and uncorrected.

No significant associations were observed for hippocampal volumes, anxiety, or affective measures (all uncorrected *ps* ≥ .12).

### Exploratory subgroup analyses

Subgroup analyses were conducted separately for tamoxifen (*n* = 8) and letrozole (*n* = 12) groups (Table 7). In the tamoxifen group, a significant increase in somatic depressive symptoms (t(7) = - 5.70, p < .001, Cohen’s d = -2.01) and in state anxiety (t(7) = 3.66, p = .008, Cohen’s d = 1.29) was observed (Table 7). The letrozole group only showed a significant decrease in positive affect (t(11) = 2.37, p =.037, Cohen’s d = .687).

**Table 7.** Paired-Samples t-Tests for Psychological Measures – exploratory subgroup analyses.

| **Tamoxifen (n=8) Measure** | **T1**<br>**Mean (SD)** | **T2**<br>**Mean (SD)** | **t** | **df** | **p** |
| --- | --- | --- | --- | --- | --- |
| BDI Cognitive–Affective | 4.75 (5.80) | 3.75 (4.83) | 0.89 | 7 | .401 |
| BDI Somatic | 3.50 (2.82) | 6.12 (3.31) | -5.70 | 7 | **<.001** |
| STAI-S | 35.87 (9.42) | 38.00 (9.70) | 3.66 | 7 | **.008** |
| PANAS Negative | 15.62 (5.23) | 14.12 (4.54) | 0.81 | 7 | .440 |
| PANAS Positive | 24.62 (3.11) | 26.25 (5.33) | -1.06 | 7 | .321 |
| **Letrozole (n=12) Measure** | **T1**<br>**Mean (SD)** | **T2**<br>**Mean (SD)** | **t** | **df** | **p** |
| BDI Cognitive–Affective | 3.66 (2.06) | 3.41 (2.99) | 0.35 | 11 | .731 |
| BDI Somatic | 4.00 (2.95) | 4.33 (3.11) | -0.63 | 11 | .540 |
| STAI-S | 38.25 (7.96) | 37.16 (5.90) | 0.84 | 11 | .418 |
| PANAS Negative | 14.08 (5.35) | 12.33 (3.55) | 1.83 | 11 | .094 |
| PANAS Positive | 25.50 (3.72) | 22.50 (4.87) | 2.37 | 11 | **.037** |

*Note.* Results are reported separately for the tamoxifen and letrozole groups. BDI: Beck’s Depression Inventory, PANAS: Positive and Negative Affect Scale; STAI-S: State-Trait Anxiety Inventory - State Anxiety Scale

With respect to structural brain changes, no significant longitudinal changes in cortical thickness were observed for either the tamoxifen (t2 > t1: pFWE-corr = .948; t1 > t2: pFWE-corr = .867) or letrozole subgroup (t2 > t1: pFWE-corr = .129; t1 > t2: pFWE-corr = .739). For gyrification, no suprathreshold clusters emerged in the letrozole group in either direction. In the tamoxifen group, the t2 > t1 contrast likewise yielded no suprathreshold clusters; the t1 > t2 contrast did not survive cluster-level FWE correction (k = 30, pFWE-corr = .228), although this cluster reached significance after FDR correction (qFDR = .040, MNI [21, −33, −5]) and should therefore be interpreted with caution.

With respect to brain–behavior associations, detailed subgroup-specific correlations are presented in Table 6b. In the letrozole group, changes in left amygdala GMV were positively associated with depressive symptoms, including total BDI-2 scores (ρ = .76, p = .004) and somatic symptoms (ρ = .80, p = .001). In contrast, no significant associations were found in the tamoxifen group (all *p*s ≥ .18).

## 4. Discussion

In this ongoing longitudinal MRI study, we examine whether the initiation of antiestrogen therapy is associated with detectable structural brain changes, targeting GMV, cortical thickness, and gyrification, and alterations in affective symptoms in women diagnosed with breast cancer (BC). The present preprint reports interim findings from the currently available sample and should therefore be interpreted as preliminary. Given that tamoxifen and letrozole differ in their mechanism of estrogen modulation, we additionally explored whether structural and affective changes varied descriptively between these two treatment subgroups. Overall, the main finding was the absence of robust macrostructural brain changes over the first 2–3 weeks following treatment initiation. We did not observe significant early volumetric changes in the hippocampus or amygdala, and no whole-brain effects survived correction for multiple comparisons at the cluster level. Regarding mental health, the strongest longitudinal finding was an increase in somatic depressive symptom scores in the BC group relative to healthy controls. Exploratory analyses further indicated a localized peak-level finding in the right anterior insula/inferior frontal operculum and uncorrected associations between changes in left amygdala volume and depressive symptoms. However, these neural findings were exploratory, were not supported by a significant time x group interaction, or did not survive correction for multiple comparisons, and should therefore be considered hypothesis-generating rather than confirmatory.

The absence of significant volume changes in the hippocampus and amygdala may partly reflect the timing of the follow-up assessment. However, structural MRI measures can change over relatively short periods, including across the menstrual cycle (53–56). The present findings therefore should not be interpreted as suggesting that detectable structural changes necessarily require several months to emerge. Rather, potential changes during the early phase of antiestrogenic therapy may have been too small or variable to be detected in hippocampal and amygdala volumes in the present sample. Limited longitudinal evidence suggests that brain structural alterations during antiestrogenic therapy become more apparent over longer time periods, for example, across the first year of treatment (33), and earlier findings on tamoxifen and estrogen-related modulation of brain structure and function point in a similar direction (32). Taken together, the absence of significant effects in our data does not necessarily indicate that neural changes are absent, but may instead reflect a combination of limited statistical power and an assessment window too short to capture macrostructural alterations; the present findings may thus reflect an early phase in which potential neural effects are still subtle or not yet detectable at the structural level, microstructural changes may precede volumetric alterations (33,57).

The insula has been consistently implicated in interoceptive awareness as well as affective and salience processing (58–60). Previous large-scale studies have reported reduced cortical thickness in the insula in individuals with major depressive disorder (61), typically assessed at a single, often unspecified stage of illness rather than shortly after symptom onset. In contrast, the present finding reflected a localized increase in cortical thickness within the BC group from t1 to t2. However, the cluster did not survive cluster-level correction, and the corresponding time x group interaction was not significant. Therefore, this result cannot be interpreted as a treatment-specific structural effect. Rather than considering the cortical thickness finding in isolation, it may be worth noting that it occurred alongside an increase in somatic depressive symptoms within the same group, although the two were analyzed separately and no direct statistical association between them was tested. If early changes in somatic symptoms indeed reflect shifts in bodily awareness, this could plausibly relate to interoceptive processes involving the insula, and such changes might emerge before broader affective alterations become apparent. This remains speculative, as it is based on the temporal co-occurrence of two independently analyzed exploratory findings rather than a demonstrated association; we offer it as a hypothesis for future studies to test directly, in line with the broader idea that early treatment-related neural effects may be more closely linked to bodily experience than to global mood changes (62–64).

An additional exploratory finding was the association between increases in left amygdala GMV and increases in depressive symptom scores. Although no group-level change in amygdala volume was detected, individuals who showed greater increases in left amygdala volume also tended to show greater increases in depressive symptoms, yet none of the participants had clinically relevant depressive symptom scores. This pattern is broadly consistent with the role of the amygdala in emotional processing, affective processing, and emotional regulation (65,66). Although, a previous meta-analysis has reported that the amygdala structure does not differ in patients with a depression diagnosis (67). Taken together, these findings suggest that early neural adaptation to antiestrogen therapy may be heterogeneous across individuals and may initially become apparent through individual brain–behavior associations rather than uniform structural changes detectable at the group level.

When examining treatment subgroups, the association between amygdala volume change and depressive symptoms appeared somewhat more pronounced in patients receiving letrozole compared to those receiving tamoxifen. This pattern may be related to the fact that aromatase inhibitors such as letrozole substantially suppress circulating estrogen levels (68), in contrast to tamoxifen, which blocks estrogen receptor activity without significantly lowering estrogen levels, a difference that could in turn influence affect-related neural systems (69,70). Since patients receiving letrozole were postmenopausal, they began treatment from a lower estrogen baseline. The extent to which further suppression from this baseline influences neural plasticity differently than the more acute estrogenic shift experienced by premenopausal tamoxifen recipients is unclear. Blood samples were collected at both time points to allow for direct assessment of estrogen and related hormone levels; these analyses are ongoing and will be reported separately. Future analyses including circulating hormone levels will be necessary to determine whether individual differences in hormonal change are associated with neural or psychological outcomes beyond treatment category alone.

At the mental health level, we did not observe broad changes across most affective domains. However, somatic depressive symptoms showed a significant time x group interaction, driven by an increase in the BC group. This pattern suggests that early mental effects are reflected in physical symptom burden, consistent with prior work distinguishing somatic and cognitive-affective components of depression (71,72). It may further indicate that early responses to treatment are initially experienced at a somatic level before broader affective changes become apparent. Previous research suggests that somatic depressive symptoms may make a distinct contribution to self-reported memory complaints, beyond cognitive/affective depressive symptoms, supporting the asymmetric worsening trajectories of both subscales seen in our study (73). Similarly, Christensen et al. (74) found that the association between nodal involvement and depressive symptoms was driven by the somatic subscale in a nationwide cohort of Danish women treated for early-stage invasive BC and assessed 3–4 months after surgery. Although the authors also advised against interpreting somatic depressive symptoms as purely treatment-driven, as overall depressive symptoms were strongly associated with comorbidity, health behaviors, and physical functioning, compared to disease or treatment-related factors. Thus, the increase may reflect early physical burden associated with the BC diagnosis and its treatment rather than a broader worsening of affective depression. Moreover, some treatment-specific differences emerged at the level of affective subdomains. Patients receiving tamoxifen showed an increase in somatic depressive symptoms and anxiety, whereas those treated with letrozole exhibited a reduction in positive affect relative to baseline. This pattern is broadly consistent with prior clinical reports linking antiestrogenic therapy to mood disturbances; for example, tamoxifen has been associated with new-onset depressive symptoms in BC patients (75) and may blunt the neuroprotective effects of estrogens (76), while aromatase inhibitors have similarly been linked to mood disturbances in case reports (36). At a mechanistic level, estradiol is known to modulate serotonergic and neurotrophic systems relevant to mood regulation (69), providing a plausible pathway through which different antiestrogenic agents could differentially impact affective symptoms. Rather than reflecting a general worsening of mood due to the diagnosis and treatment, these findings point to more selective changes in specific affective dimensions.

This stability in broader affective measures may reflect either a true absence of short-term changes or limited sensitivity to detect subtle alterations within the relatively short observation period. At the same time, somatic symptoms, but not broader affective measures, showed significant change, suggesting that early responses to treatment may be selective rather than global. More broadly, previous studies on the cognitive and affective effects of antiestrogenic therapy have reported heterogeneous findings, likely reflecting differences in treatment type, duration, and patient characteristics (77).

Taken together, our findings point to a temporal dissociation between mental and neuroanatomical changes in the early phase of antiestrogen therapy: measurable shifts in somatic depressive symptoms and affect emerged within the first weeks of treatment initiation, whereas cortical and subcortical structure remained largely stable. This suggests that, at least in this early window, behavioral and psychological measures may be more sensitive indicators of treatment-related change than structural MRI metrics, which may require longer observation periods to reveal detectable alterations.

It is also worth noting that much of the existing neuroimaging research in BC has focused on chemotherapy-related effects, as reflected in a recent systematic review and meta-analysis on chemotherapy-related cognitive impairment (*chemobrain*) in BC patients and a broader review of the cognitive effects of cancer and cancer treatments (78,79), whereas considerably less is known about the early neural impact of antiestrogenic therapy specifically. In this context, our findings suggest that early effects, if present, are likely subtle and not easily captured by standard group-level structural analyses.

## Limitations

Several limitations should be considered. First, this is an ongoing study, thus the relatively small sample size which limits statistical power, particularly for subgroup analyses, at the moment. Second, multiple statistical tests were conducted across mental and neuroimaging outcomes without formal correction in all cases, increasing the risk of type I error. Therefore, findings, especially those at trend or exploratory levels, should be interpreted with caution.

## Conclusion

The preliminary results reported here indicate that no robust group-level macrostructural brain changes were detected within the first 2–3 weeks following the initiation of antiestrogen therapy in women with BC. However, the absence of robust effects in this interim sample does not exclude the possibility of early structural changes, which may be subtle, heterogeneous, or difficult to detect at the group level. Exploratory findings, including a localized increase in cortical thickness in the right anterior insula/inferior frontal operculum and positive associations between changes in left amygdala volume and depressive symptoms, may point to early neural correlates but should be interpreted cautiously. At the same time, somatic depressive symptoms increased during this early treatment period, indicating that psychological changes may already be evident within the first weeks of treatment. Larger samples and longer follow-up periods are needed to determine the timing and magnitude of structural brain and psychological changes during antiestrogen therapy.

## CRediT authorship contribution statement

**Serenay Yazici Sarikaya:** Conceptualization, Data curation, Formal analysis, Investigation, Resources, Methodology, Recruitment, Software, Validation, Writing – original draft, Writing – review & editing. **Berfin Gülbahçe:** Software, Recruitment, Visualization, Writing – review & editing. **Ann-Christin S. Kimmig:** Conceptualization, Validation, Writing – review & editing. **Markus Hahn:** Recruitment, Validation, Writing – review & editing. **Sara Y. Brucker:** Conceptualization, Funding acquisition, Recruitment, Writing – review & editing. **Benjamin Bender:** Conceptualization, Writing – review & editing. **Uta Hoopmann:** Conceptualization, Writing – review & editing. **Anna Wikman:** Conceptualization, Funding acquisition, Methodology, Resources, Supervision, Validation, Writing – review & editing. **Birgit Derntl:** Conceptualization, Funding acquisition, Methodology, Project administration, Resources, Supervision, Validation, Writing – review & editing.

## Data Availability

All data produced in the present study are available upon reasonable request to the authors

## Acknowledgments

The authors would like to thank all individuals who supported data collection and participant recruitment (medical students: Luise Pfleiderer, Shona Ramroth, Emma Karrlein, Emily Luedicke, Johanna Baltes, and Melike Mermer).

## Availability of data and materials

The data supporting the findings of this study are available from the corresponding author upon reasonable request.

